# *CREBRF* missense variant rs373863828 has both direct and indirect effects on type 2 diabetes and fasting glucose in Polynesians living in Samoa and Aotearoa New Zealand

**DOI:** 10.1101/2021.02.15.21251768

**Authors:** Emily M. Russell, Jenna C. Carlson, Mohanraj Krishnan, Nicola L. Hawley, Guangyun Sun, Hong Cheng, Take Naseri, Muagututi‘a Sefuiva Reupena, Satupa‘itea Viali, John Tuitele, Tanya J. Major, Iva Miljkovic, Tony R. Merriman, Ranjan Deka, Daniel E. Weeks, Stephen T. McGarvey, Ryan L. Minster

## Abstract

**Objective:** The minor allele of rs373863828 in *CREBRF* is associated with higher BMI, lower fasting glucose, and lower odds of type 2 diabetes. We examined the associations between BMI and rs373863828 on type 2 diabetes and fasting glucose with a large sample of adult Polynesians from Samoa, American Samoa and Aotearoa New Zealand and estimated direct and indirect (via BMI) effects of rs373863828 on type 2 diabetes and fasting glucose.

**Research Design and Methods:** We regressed type 2 diabetes and fasting glucose on BMI and rs373863828 stratified by obesity, regressed type 2 diabetes and fasting glucose on BMI stratified by rs373863828 genotype, and assessed the effects of rs373863828 on type 2 diabetes and fasting glucose with path analysis.

**Results:** Association of BMI with fasting glucose was greater in those without obesity than in those with obesity. We did not observe evidence of differences by genotype. In the path analysis, the minor allele has direct negative and indirect positive effects on type 2 diabetes risk and fasting glucose, with the indirect effect mediated through a *direct* positive effect on BMI.

**Conclusions:** There may be a stronger effect of BMI on fasting glucose in Polynesians without obesity than in those with obesity. Carrying the rs373863828 minor allele does not decouple higher BMI from odds of type 2 diabetes. Given the current cost of genotyping compared to the accessibility of measuring BMI, including rs373863828 as a clinical predictor of type 2 diabetes may not be indicated.

## Main Document

High BMI is a risk factor for type 2 diabetes and elevated fasting glucose levels. While type 2 diabetes and obesity are highly correlated, there is little overlap in genetic variants associated with each phenotype [1]. One genetic variant associated with both, the minor allele A of a missense variant in *CREBRF* (rs373863828, c.1370G>A, p.R457Q) is simultaneously associated with higher BMI (1.36 kg/m^2^ per copy of the A allele) and, contrary to expectations, lower odds of type 2 diabetes (0.586 OR) and lower fasting glucose (−1.65 mg/dL per copy of the A allele) in Samoans and other Pacific Island populations [2–6]. Alleles with such discrepant effects have been termed “favorable adiposity” alleles [7, 8]. Favorable adiposity alleles like those of rs373863828 in *CREBRF* and variants in *ADAMTS9, GRB14/COBLL1*, and *TCF7L2* [1, 9] could reveal avenues for the study of the variation in biological pathways underlying the relationship between BMI and type 2 diabetes. The biological mechanisms of *CREBRF* and its role in development of obesity and type 2 diabetes remain largely unknown.

The association of rs373863828 with BMI, obesity, type 2 diabetes, and fasting glucose was first identified in Samoans from Samoa and American Samoa [2] and has since been replicated in other Pacific populations[3–6]. rs373863828 is common in Samoans (minor allele frequency [MAF] = 0.259)[2], Tongans (MAF = 0.150)[5], Pukapukans (MAF = 0.243), Niueans (MAF = 0.096), Cook Island Māori (MAF = 0.195), New Zealand Māori (MAF = 0.174)[3], Native Hawaiians (MAF = 0.128)[6], and Pacific Island people from Guam and Saipan (MAF = 0.042)[4] but is very rare in non-Pacific populations (MAF < 0.001) [10, 11].

The prevalence of obesity in Polynesia is among the highest in the world, and temporal increases in obesity-related disorders such as type 2 diabetes represent a growing public health problem in the region [12]. From 1978 to 2013, the prevalence of obesity (≥ 30 kg/m^2^) in men rose from 28% to 53% and in women, from 45% to 78% [13]. Similarly, from 1980 to 2014, the age-standardized prevalence of type 2 diabetes of Samoan men and women increased from 6.1% and 9.0%, respectively, to 22.7% and 26.6%, respectively [14].

Previous work has focused on the direct associations of rs373863828 with BMI, type 2 diabetes, and fasting glucose [2–5]. Here we examine, more extensively, the relationships among type 2 diabetes, fasting glucose, BMI, and rs373863828 in three samples of adult Samoans (n = 2,861, n = 1,083 and n = 1,013) living in Samoa and American Samoa and a sample of adult Polynesians (n = 1,270) of several ancestries living in Aotearoa New Zealand. This combined study (n = 7,127) is the largest one thus far examining the relationship between rs373863828, type 2 diabetes, and BMI [2, 3]. This is also the first time that path analysis has been used to explore the direct and indirect effects of rs373863828 on type 2 diabetes and fasting glucose. Studies such as this may inform the utility of including a screen for rs373863828 in clinical practice.

## Methods

### Participants and Phenotypes

For this study, we worked with three samples of Samoan participants from Samoa and American Samoa [2, 15–20] from a 1990–95 longitudinal study of cardiometabolic disease, a 2002–03 family-based cross-sectional study of adiposity-related trait genetics, and a 2010 population-based study of cardiometabolic disease genetics; and a sample of participants of Māori and Pacific (Polynesian) ancestry from the Genetics of Gout, Diabetes and Kidney Disease in Aotearoa New Zealand study [3].

Adult participants in the longitudinal 1990–95 study were recruited from Samoa and American Samoa. They reported Samoan ancestry and no previous diagnosis of diabetes, heart disease, or hypertension at baseline [18–20]. Whole blood was collected after an overnight fast for the fasting glucose measurement. Fasting glucose ≥ 126 mg/dl at baseline was used to define type 2 diabetes [2, 18]. The final sample was comprised of 1,013 individuals with full phenotypic and genotypic data (Table 1a).

**Table 1:**
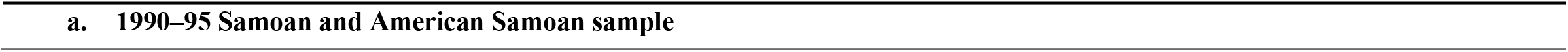
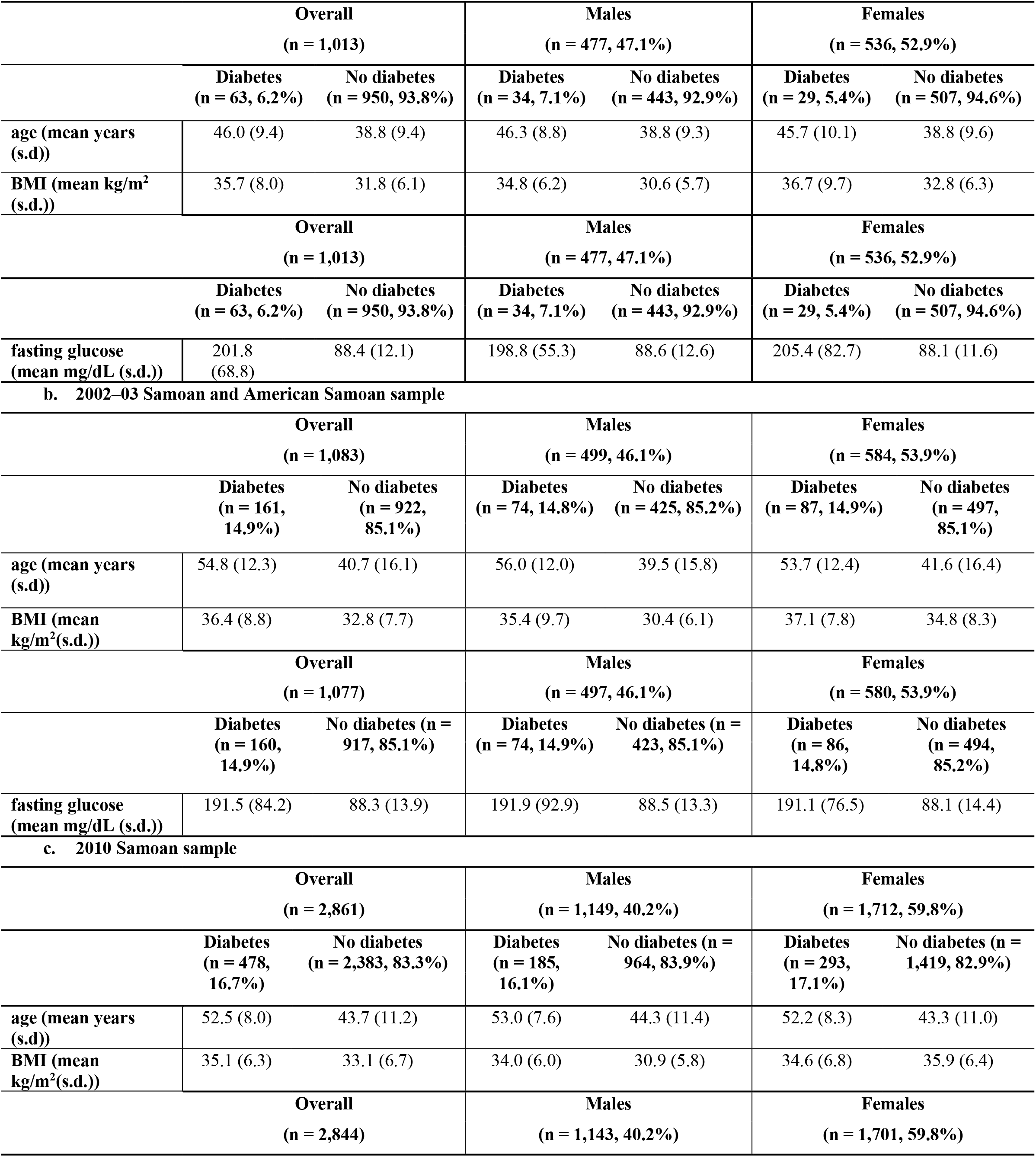
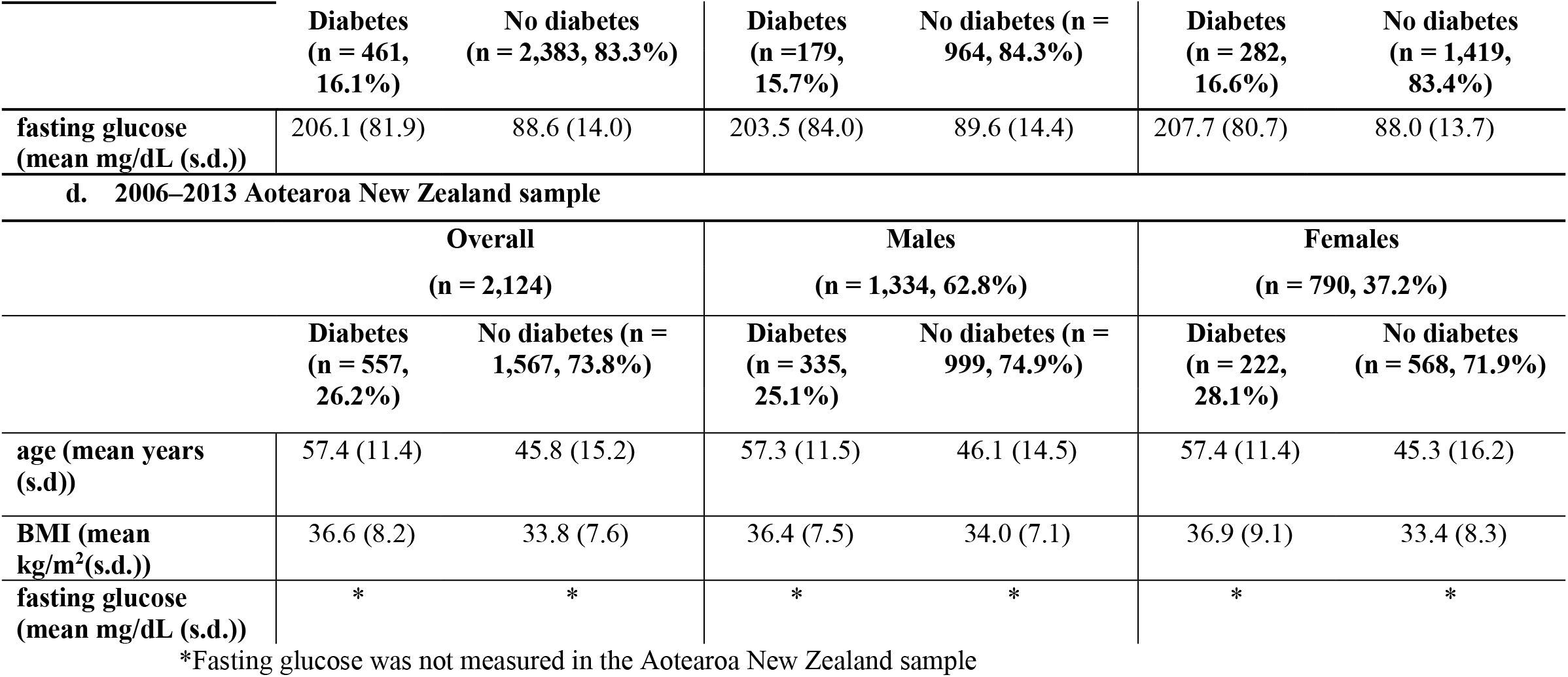
Sample demographics information stratified by sex and diabetes status

Participants of the cross-sectional 2002–03 family study reported Samoan ancestry and were recruited from Samoa and American Samoa as members of families of probands selected from the longitudinal 1990–1995 study [2, 16, 17]. Whole blood was collected after a 10 hour fast for the fasting glucose measurement. Fasting glucose ≥ 126 mg/dl and/or a previous diabetes diagnosis was used to define type 2 diabetes [2, 16]. The final sample was comprised of 1,083 adults with full phenotypic and genotypic data (Table 1b).

Participants in the 2010 Samoan sample were drawn from a cross-sectional population-based study of adults of reported Samoan ancestry recruited in Samoa. Details of the sample population and recruitment have been described elsewhere [2, 15]. In brief, whole blood samples were collected following a 10 hour fast for the fasting glucose measurement. Type 2 diabetes was defined as fasting glucose ≥ 126 mg/dl and/or currently taking medication to treat diabetes [2]. The final sample was comprised of 2,861 participants with full phenotypic and genotypic data (Table 1c).

The sample from Aotearoa New Zealand (n=2,124) was recruited from 2006**–**2017 [21]. Type 2 diabetes was ascertained by physician-diagnosis and/or participant reports and/or use of glucose lowering therapy [3]. Ancestry of participants was based on both self-reported NZ Māori and/or Pacific ancestries of their grandparents and clustering of genome-wide principal components resulting in 2,124 adults of Polynesian ancestry. Fasting glucose was not measured in this study (Table 1d).

In all studies, height and weight were measured in duplicate and were used to calculate BMI. Compared to people of European ancestries, people of Polynesian ancestries have a greater ratio of lean mass to fat mass at higher BMIs [22, 23]. Therefore, when classifying participants into those with and without obesity, we used a threshold of BMI > 32 kg/m^2^, which is more appropriate in this population than the standard WHO cutoff of ≥ 30 kg/m^2^ [22].

Details of the DNA extraction and genotyping for the Samoan, American Samoan and Aotearoa New Zealand samples have been described elsewhere [2, 3]. Subject relatedness in the 2010 sample was measured using an empirical kinship matrix calculated with OpenMendel using genome-wide genotype data [2, 24, 25]. In the 2002–03 sample, expected kinship was derived from familial pedigrees with OpenMendel [24]. Neither genome-wide genotype data nor pedigree information was available for participants in the 1990–95 sample, so subjects were treated as unrelated in statistical models. In the Aotearoa New Zealand sample, a kinship coefficient matrix was calculated from 40,156 independent autosomal markers using PLINK v1.9 [26].

All participants in these studies gave written informed consent. The research in Samoa and American Samoa was reviewed and approved by the institutional review boards of Miriam Hospital, Providence, RI; Brown University; University of Cincinnati; and University of Pittsburgh. Research in Samoa was also reviewed and approved by the Health Research Committee of the Samoan Ministry of Health. Research in American Samoa was additionally reviewed and approved by the American Samoa Department of Health institutional review board. Ethical approval for the Aotearoa New Zealand study was given by the NZ Multi-Region Ethics Committee (MEC/05/10/130; MEC/10/09/092; MEC/11/04/036) and the Northern Y Region Health Research Ethics Committee (NPHCT study; NTY07/07/074).

### Statistical Analysis

We explored the impact of BMI and rs373863828 on type 2 diabetes and fasting glucose in three ways: (1) regression analysis to assess the effect of rs373863828 and natural log transformed BMI (lnBMI) on type 2 diabetes and natural log transformed fasting glucose (lnFG) in the two obesity-stratified groups; (2) regression analysis to assess the effect of lnBMI on type 2 diabetes and lnFG within each of the three rs373863828 genotype groups; and (3) path analysis to model the effects of lnBMI and rs373863828 on type 2 diabetes and lnFG.

Except where noted, all statistical analyses were conducted using R (version 3.6.0), and a significance threshold of α < 0.05 was used for all tests [27]. Age and age^2^ were scaled and centered (cAge and cAge^2^), to separate the effects of the sex × age and sex × age^2^ interactions from the main effects. The rs373863828 genotype was coded as an additive genetic effect so that it represents the number of copies of the minor allele, A. Participants who reported a diabetes diagnosis—and thus were potentially being treated for the disease—were excluded from fasting glucose analyses (Table 1). We did not exclude individuals with fasting glucose ≥ 126 mg/dL from the fasting glucose analysis if they did not report a diagnosis because they were presumed to not be part of any intervention affecting their fasting glucose levels.

Each regression model described below was fit using logistic mixed models for type 2 diabetes status and linear mixed models for fasting glucose (Table S1). We adjusted for kinship as a random effect using the *lme4qtl* package [28].

Unless otherwise noted, we performed each analysis in each of the three Samoan/American Samoan samples and the Aotearoa New Zealand sample and then meta-analyzed the results together. In the regression analyses, the samples were meta-analyzed using standard error–weights based on the effect sizes and heterogeneity was tested in METAL [29]. In the path analyses, we pooled the raw data across the samples.

All regression models below were adjusted for the following fixed-effect covariates: cAge, cAge^2^, sex, sex × cAge, sex × cAge^2^. The analyses in the 2002–03 and 1990–95 Samoan/American Samoan sample sets were also adjusted for polity (Samoa or American Samoa) to account for different economic and cultural contexts. For each regression model, we compared the effect sizes and the overlap of confidence intervals, a conservative measure of heterogeneity, to assess the whether the effects were different per stratification [30].

### Association of lnBMI and rs373863828 with type 2 diabetes and lnFG stratified by obesity

We stratified the samples by the Polynesian obesity cutoff (32 kg/m^2^). We examined if the effects of lnBMI and rs373863828 on type 2 diabetes differed for individuals with and without obesity by regressing type 2 diabetes on lnBMI and rs373863828 in the obesity-stratified groups. We also examined whether the effects of lnBMI and rs373863828 on fasting glucose differed by regressing lnFG on lnBMI and rs373863828 in the obesity-stratified groups.

### Association of lnBMI with type 2 diabetes and lnFG stratified by rs373863828 genotype

We stratified the samples by rs373863828 genotype (GG, GA, and AA). We examined if the effect of lnBMI on type 2 diabetes differed depending on genotype group by regressing type 2 diabetes on lnBMI in each of the groups. We also examined whether the effect of lnBMI on lnFG differed by regressing lnFG on BMI in each of the genotype groups.

### Modeling the effects of lnBMI and rs373863828 on type 2 diabetes and lnFG with path analysis

The relationships among sex, age, rs373863828, lnBMI, and type 2 diabetes were modeled with path analysis using the *lavaan* package in R [31]. Interaction terms were not included in the path model, and therefore variables were not centered. In the path analysis model, we regressed lnBMI on sex, age, and rs373863828, and type 2 diabetes on lnBMI, sex, age, and rs373863828. The indirect effect of rs373863828 on type 2 diabetes as mediated by lnBMI was calculated by multiplying the path coefficient of the direct effect of rs373863828 on lnBMI by the path coefficient of the direct effect of lnBMI on type 2 diabetes. Fasting glucose was modeled using path analysis by substituting lnFG for type 2 diabetes in the above model. Relatedness was not accounted for in these models because the methods for accounting for relatedness do not exist, and this may increase type I error.

In path analysis, the absolute value of an effect size of around 0.1 is commonly considered to be small, around 0.3 is medium, and 0.5 is large [32]. The direct paths of sex to type 2 diabetes and sex to lnFG had effect sizes less than 0.1 and were not part of the direct or indirect path of rs373863828 on type 2 diabetes/lnFG so they were trimmed from the models. For both type 2 diabetes and lnFG, two models were examined: with and without a dummy variable for study (2010 Samoa, 2002–2003 Samoa, 2002–2003 American Samoa, 1990–95 Samoa, 1990–95 American Samoa or 2006–2013 Aotearoa New Zealand) to assess for study sample differences.

## Results

### Association of lnBMI and rs373863828 with type 2 diabetes and lnFG stratified by obesity

In the regression of type 2 diabetes on lnBMI and rs373863828, higher lnBMI was associated with higher type 2 diabetes odds in participants without and with obesity (OR = 6.27, *p* = 0.003; and OR = 4.41, *p* = 3.68 × 10^−6^, respectively). lnBMI was also associated with lnFG in participants without and with obesity (β = 0.191 (1.21 mg/dL higher fasting glucose with each higher unit of lnBMI), *p =* 4.11 × 10^−6^ and β = 0.093 (1.10 mg/dL higher fasting glucose), *p =* 0.036, respectively) (Figures 1 and S1 and Table 2). There was evidence of heterogeneity (*p* < 0.01) between studies in the subset with obesity in the type 2 diabetes analysis (*p* = 0.082) and the subset without obesity in the lnFG analysis (*p* = 0.063) (Figure S1 and Table S2).

**Table 2:** Effect of lnBMI and rs373863828 on type 2 diabetes and fasting glucose in the meta-analysis, stratified by obesity status (a) and genotype (b)

| Meta-analysis<br>lnBMI |  |  |  | Meta-analysis<br>rs373863828 |  |  |
| --- | --- | --- | --- | --- | --- | --- |
| Dependent variable: type 2 diabetes |  |  |  |  |  |  |
| Stratum* | n | OR (95% CI) | *p* | n | OR (95% CI) | *p* |
| without obesity | 3,211<br>(393 cases) | 6.27<br>(1.90, 20.75) | **0.003** | 3,211 | 0.58<br>(0.45, 0.74) | **9.70 × 10<sup>-6</sup>** |
| with obesity | 3,870<br>(866 cases) | 4.41<br>(2.35, 8.25) | **3.68 × 10<sup>-6</sup>** | 3,870 | 0.73<br>(0.63, 0.85) | **2.82 × 10<sup>-5</sup>** |
| Dependent variable: natural log transformed fasting glucose |  |  |  |  |  |  |
| Stratum* | n | β (s.e.) | p | n | β (s.e.) | p |
| without obesity | 2,180 | 0.191 (0.041) | 4.11 × 10<sup>-6</sup> | 2,180 | -0.030 (0.008) | 8.17 × 10<sup>-5</sup> |
| with obesity | 2,352 | 0.093 (0.044) | 0.036 | 2,352 | -0.033 (0.009) | 1.22 × 10<sup>-4</sup> |
| *without obesity: BMI ≤ 32 kg/m<sup>2</sup>; with obesity: BMI > 32 kg/m<sup>2</sup> |  |  |  |  |  |  |

b.
| Meta-analysis<br>lnBMI |  |  |  |
| --- | --- | --- | --- |
| Dependent variable: type 2 diabetes |  |  |  |
| Stratum | n | OR (95% CI) | *p* |
| GG | 4,111<br>(824 cases) | 6.37<br>(4.03, 10.05) | $1.88 \times 10^{-15}$ |
| GA | 2,547<br>(389 cases) | 7.049<br>(2.85, 17.47) | $2.46 \times 10^{-5}$ |
| AA | 232<br>(22 cases) | 1.774<br>(0.19, 16.93) | 0.619 |

| Dependent variable: natural log transformed fasting glucose |  |  |  |
| --- | --- | --- | --- |
| Stratum | n | $\beta$ (s.e.) | *p* |
| GG | 2,442 | 0.245 (0.027) | $3.12 \times 10^{-20}$ |
| GA | 1,759 | 0.214 (0.034) | $1.89 \times 10^{-10}$ |
| AA | 331 | 0.161 (0.062) | **0.001** |

**Figure 1:**
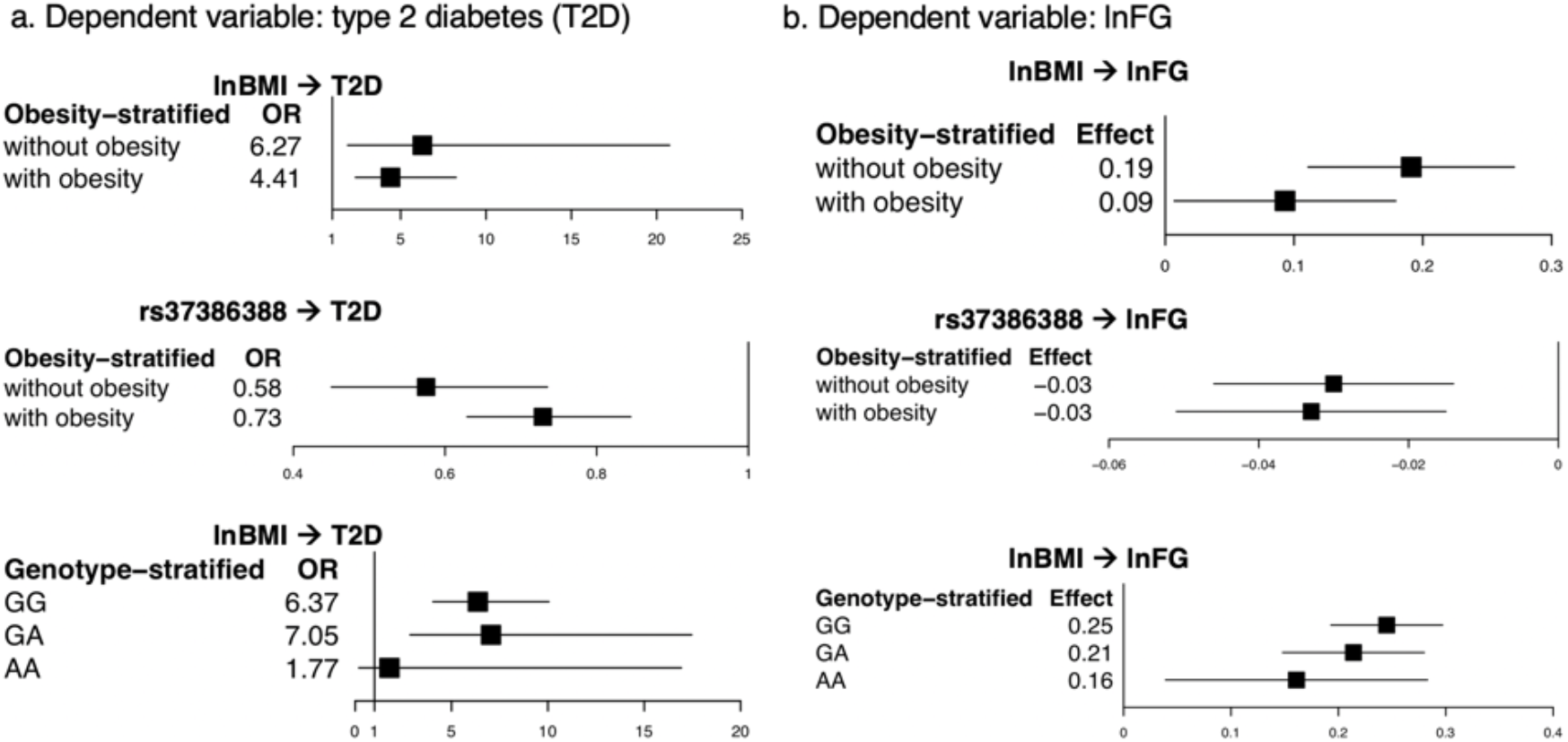
Comparison of the odds ratios and effect sizes in the obesity and genotype-stratified groups. From the meta-analysis, (a) a comparison of the odds ratios (OR) and 95% confidence intervals in the obesity and genotype-stratified models with type 2 diabetes as the independent variable and (b) a comparison of the effect sizes and 95% confidence intervals in the obesity and genotype-stratified models with lnFG as the independent variable.

On the other hand, rs373863828 was associated with lower type 2 diabetes odds in participants, both without and with obesity (OR = 0.58, *p* = 9.70 × 10^−6^; and OR = 0.73, *p* = 2.82 × 10^−5^, respectively). rs373863828 was also associated with lower lnFG in participants both without and with obesity (β = −0.030 (−0.97 mg/dL fasting glucose with each copy of the missense allele), *p* = 8.17 × 10^−5^ and β = −0.033 (−0.97 mg/dL fasting glucose), *p* = 1.22 × 10^−4^, respectively) (Figure 1 and S1 and Table 2).

### Association of lnBMI with type 2 diabetes and lnFG stratified by rs373863828 genotype

In the regression of type 2 diabetes on lnBMI in the genotype-stratified groups, higher lnBMI was associated with higher type 2 diabetes odds in both the GG and GA groups (OR = 6.37, *p* = 1.88 × 10^−15^; and OR = 7.05, *p* = 2.46 × 10^−5^, respectively). lnBMI was not significantly associated with type 2 diabetes in the AA group (OR = 1.774, *p* = 0.619) (Table 2). However, the analysis of the AA group only included participants from the 2010 Samoan sample (type 2 diabetes cases = 22) because the regression models did not converge with the low numbers of type 2 diabetes cases in the AA group in the 1990–95 and 2002–03 Samoan samples (n = 2 and n = 6, respectively) and in the Aotearoa New Zealand sample (n = 16) (Figure 1 and Table S3). According to a post-hoc power calculation, we had 70.5% power to detect an odds ratio of 1.774 in our AA sample with α = 0.05.

In the meta-analysis of lnBMI on lnFG in the genotype-stratified groups, higher lnBMI was associated with higher lnFG in all genotype groups (GG: β = 0.245 (1.28 mg/dL higher fasting glucose with each unit of lnBMI), *p* = 3.12 × 10^−20^; GA: β = 0.214 (1.24 mg/dL higher fasting glucose), *p* = 1.89 × 10^−10^; and AA: β = 0.161 (1.17 mg/dL higher fasting glucose), *p* = 0.001) (Figure 1 and Table S3). In the lnFG analyses, there was evidence of heterogeneity between studies in the GG group (*p* = 0.079) (Figure S2).

### Path analysis models

The type 2 diabetes path analysis model included five nodes: lnBMI, sex, rs373863828 genotype (subscript Gt in the following path coefficients), age, and type 2 diabetes (subscript T2D). The path pooled analyses were modeled with and without additional nodes to account for study sample (Figure S2). These models were similar so the simpler models are presented below. In the path pooled analysis displayed in Figure 2a, rs373863828 had a small direct effect on lnBMI (*P*_lnBMI,Gt_ = 0.10, *p* < 0.001). rs373863828 and lnBMI had small direct effects on type 2 diabetes (*P*_T2D,Gt_ = −0.09, *p* < 0.001; and *P*_T2D,lnBMI_ = 0.13, *p* < 0.001, respectively). rs373863828 also had a small indirect effect on type 2 diabetes as mediated by lnBMI (*P*_lnBMI,Gt_ × *P*_T2D,lnBMI_ = 0.01). The root mean square error of approximation (RMSEA) was 0.011 (90% CI 0.000–0.035) indicating a good model fit [33].

**Figure 2:**
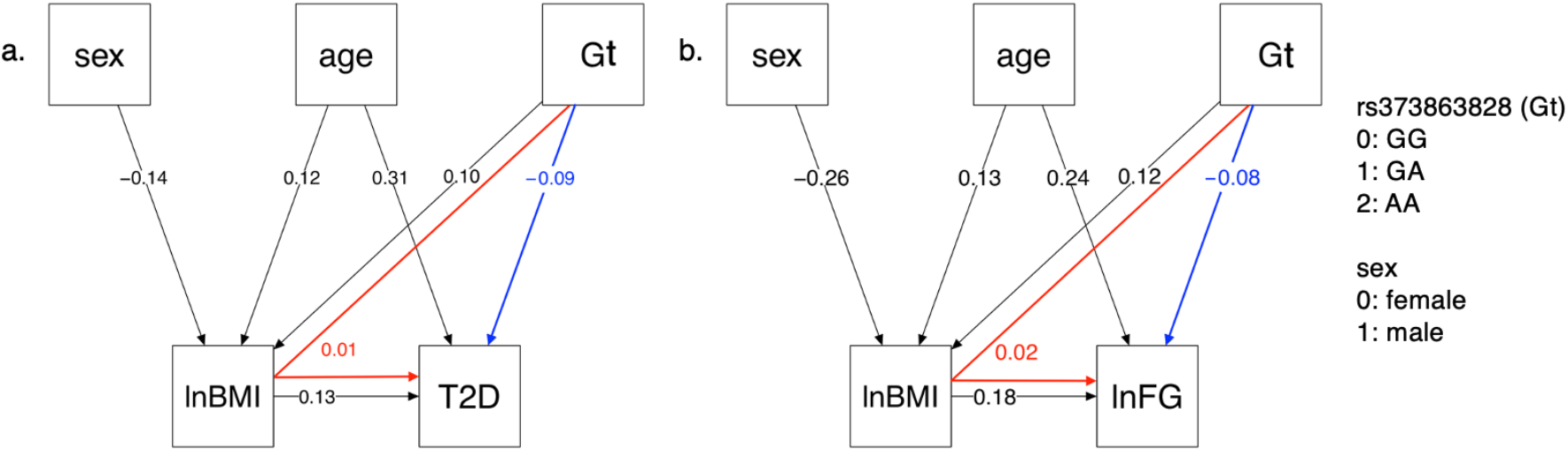
Type 2 Diabetes and lnFG Path Analysis Models. In the pooled analysis, rs373863828 genotype (Gt) has both direct (blue) and indirect (red) effects on (a) type 2 diabetes (T2D) and (b) lnFG as mediated by lnBMI. All direct paths shown have p < 0.001.

The lnFG path analysis model included five nodes: lnBMI, sex, rs373863828, age, and lnFG. In the path pooled analysis (Figure 2b), rs373863828 had a small direct effect on lnBMI (*P*_lnBMI,Gt_ = 0.12, *p* < 0.001). rs373863828 had a small direct effect on lnFG (*P*_lnFG,lnBMI_ = 0.18, *p* < 0.001). rs373863828 also had a small indirect effect on lnFG that was mediated by lnBMI (*P*_lnBMI,Gt_ × *P*_lnFG,lnBMI_ = 0.02). RMSEA was 0.068 (90% CI 0.045–0.094) indicating that the path model was a mediocre fit [33].

## Discussion

We see here that the discordant association of rs373863828 with BMI and type 2 diabetes is consistent among individuals with and without obesity and across dietary and physical activity exposures and general nutritional environments from Samoa in 1991 [19] to Aotearoa New Zealand in 2006–13 [3, 34]. While there is evidence of heterogeneity between studies in some of the meta-analyses, this is not surprising, giving the secular trends in fasting glucose and type 2 diabetes. These samples were recruited during different decades and from different countries, and there could be underlying differences in them arising from cohort effects, their different dietary and physical-activity environments and differences in other exposures. This heterogeneity may limit our power to detect associations.

There may be a stronger effect of BMI on fasting glucose levels in Samoans without obesity than in Samoans with obesity, especially in the more recent samples. This is likely attributable to more rapid increases in hyperglycemia and insulin resistance as adiposity increases from normal to overweight to obesity, relative to later in the pathophysiology process [35]. While higher BMI is associated with higher fasting glucose across all rs373863828 genotypes, the magnitude of effect is less with each copy of the missense allele. However, we do not see a similar pattern in the effect of BMI on type 2 diabetes. Carrying the A allele does not decouple higher BMI from higher odds of type 2 diabetes. Individuals who are GG or GA or AA all have higher odds of type 2 diabetes with higher BMI, but individuals who are AA have a lower risk of type 2 diabetes than those who are GG at any given BMI.

In our exploration of the relationships among BMI, fasting glucose, and genotype via path analysis, we found that the rs373863828 missense variant has both a *direct* protective effect on type 2 diabetes and fasting glucose and an *indirect* opposing risk-increasing effect on type 2 diabetes odds and fasting glucose. This indirect effect is mediated through the direct increasing effect of the rs373863828 missense variant on BMI. Again, the rs373863828 missense allele provides some protection from type 2 diabetes, but higher BMI is associated with higher type 2 diabetes odds regardless of rs373863828 genotype.

Little is known about the biology behind the effects of *CREBRF* on type 2 diabetes and BMI. There is evidence that it has multiple, tissue-specific function. The rs373863828 minor allele is associated with greater bone and lean mass in Samoan infants at four months old, but it is not associated with BMI [36], suggesting that body composition may be involved in the association between rs373863828, type 2 diabetes, and BMI. Another function is indicated by the analog of *CREBRF* in *Drosophila* which is involved in energy homeostasis [37], and along with one of its binding partners, CREBL2, it functions as a metabolic regulator linking nutrient sensor mTORC1 to cellular metabolic response [38]. Finally, there is evidence of a role in adipose differentiation. Overexpression of CREBRF in mouse 3T3-L1 preadipocytes induces the expression of adipogenic markers and results in increased lipid accumulation, and overexpression of *CREBRF*:p.R457Q promotes greater lipid storage while using less energy than wild-type *CREBRF* [2]. Our path analysis with its disparate effects adds to this body of evidence that *CREBRF*’s role in biology is multifarious.

While rs373863828 has disparate effects on BMI and type 2 diabetes, BMI is still an important and readily available clinical predictor of type 2 diabetes risk [39], independent of rs373863828. Currently, given the high cost of genotyping and ease of measuring BMI across contexts, there is limited utility to including genetic factors in risk prediction models for type 2 diabetes [1, 40]. While this work is unlikely to impact clinical practice in the immediate future, exploring the effect of rs373863828 on BMI and type 2 diabetes adds to current knowledge of the effects of *CREBRF* on BMI and type 2 diabetes.

In summary, we provide evidence that the rs373863828 minor allele has both a direct negative effect and an indirect positive effect on type 2 diabetes and fasting glucose, respectively. We also suggest that there may be a stronger positive association between BMI and type 2 diabetes and fasting glucose in Polynesians without obesity than in Polynesians with obesity. Finally, the A allele of rs373863828 is associated with lower odds of type 2 diabetes, but no matter the genotype, higher BMI is still associated with higher odds of type 2 diabetes.

## Data Availability

The 2010 sample data from Samoa examined in this study are available from dbGaP (accession number: phs000914.v1.p1). The 1991-95 and 2002-03 sample data from Samoa and American Samoa is not available because participants did not consent to data sharing when enrolled. Sample data from Aotearoa New Zealand is not publicly available owing to consent restrictions but can be requested from author TRM under an appropriate arrangement.

## Data Availability

The 2010 sample data from Samoa examined in this study are available from dbGaP (accession number: phs000914.v1.p1). The 1991–95 and 2002–03 sample data from Samoa and American Samoa is not available because participants did not consent to data sharing when enrolled. Sample data from Aotearoa New Zealand is not publicly available owing to consent restrictions but can be requested from author TRM under an appropriate arrangement.

## Acknowledgements

We are grateful for the contributions of Melania Selu and Vaimoana Lupematisila to the recruitment of the 2010 Samoan sample. The authors thank the Samoan participants and local village authorities and research assistants over the years. We acknowledge the support of our research collaboration with the Samoa Ministry of Health, Samoa Bureau of Statistics, Samoan Ministry of Women, Community and Social Development and the American Samoa Department of Health.

The authors acknowledge the contributions of Nicola Dalbeth, Janak de Zoysa, Rinki Murphy and Lisa Stamp to the recruiting of the Aoteaora New Zealand sample, Jennie Harré Hindmarsh to the recruiting of the Ngati Porou Hauora (NPHCT) sample included within the Aoteaora New Zealand sample, and the contributions of Ruth Topless, Amanda Phipps-Green, Marilyn Merriman and Murray Cadzow for laboratory work and management of the New Zealand datasets.

## Contribution statement

EMR and MK performed the analyses with guidance from RLM, DEW, JCC, and IM. EMR, JCC, and MK wrote the manuscript. NLH led the field work data collection in Samoa and phenotype analyses with guidance from STM. GS and HC performed the genotyping of the Samoan samples with guidance from RD. MSR, SV, and JT facilitated fieldwork in Samoa and American Samoa. TN contributed to the discussion of the public health implications of the findings. TRM contributed to the data collection in Aotearoa New Zealand. TJM created the kinship matrix for the Aoteaora New Zealand sample. All authors contributed to this work, discussed the results, and critically reviewed and revised the manuscript. RLM is the guarantor of this work.

## Abbreviations

cAge: mean-centered scaled age
cAge2: mean-centered scaled age squared
*CREBRF*: cAMP-responsive element binding protein 3 regulatory factor
lnBMI: natural logarithm of BMI
lnFG: natural logarithm of fasting blood glucose
MAF: minor allele frequency
OR: odds ratio
RMSEA: root mean square error of approximation

