## Supplemental Tables and Figures for "*CREBRF* missense variant rs373863828 has both direct and indirect effects on type 2 diabetes and fasting glucose in Polynesians living in Samoa and Aotearoa New Zealand"

**Supplementary Table S1:** Equations for the linear mixed models

| Model | Equation |
|---|---|
| obesity-stratified | T2D ~ (cAge × sex) + ($cAge^2$ × sex) + cAge + sex + $cAge^2$ + gt + lnBMI + k<br><br>lnFG ~ (cAge × sex) + ($cAge^2$ × sex) + cAge + sex + $cAge^2$ + gt + lnBMI + k |
| genotype-stratified | T2D ~ (cAge × sex) + ($cAge^2$ × sex) + cAge + sex + $cAge^2$ + lnBMI + k<br><br>lnFG ~ (cAge × sex) + ($cAge^2$ × sex) + cAge + sex + $cAge^2$ + lnBMI + k |

T2D = type 2 diabetes, k = kinship random effect, gt = additive rs373863828 genotype, cAge = centered and scaled age

**Supplementary Table S2:** Effect of (a) lnBMI and (b) rs37386388 on type 2 diabetes and fasting glucose in each cohort, stratified by obesity status

a. Predictor: lnBMI

| | 1990–95 Samoan cohort lnBMI | | | 2002–03 Samoan cohort lnBMI | | | 2010 Samoan cohort lnBMI | | | 2006–2013 Aotearoa New Zealand cohort lnBMI | | |
|---|---|---|---|---|---|---|---|---|---|---|---|---|
| **Dependent variable: type 2 diabetes** | | | | | | | | | | | | |
| **Stratum *** | **n** | **OR (95% CI)** | ***p*** | **n** | **OR (95% CI)** | ***p*** | **n** | **OR (95% CI)** | ***p*** | **n** | **OR (95% CI)** | ***p*** |
| without obesity | 546 | 0.194 (0.004, 10.430) | 0.419 | 527 | 16.275 ($1.65 \times 10^{-5}$, $1.61 \times 10^{7}$) | 0.692 | 1,265 | 12.692 (2.141, 75.240) | **0.005** | 873 | 6.104 (1.028, 36.257) | **0.047** |
| with obesity | 467 | 16.676 (1.411, 197.078) | **0.026** | 556 | 6.032 (1.158, 31.416) | **0.033** | 1,596 | 1.484 (0.515, 4.278) | 0.465 | 1,251 | 7.807 (3.023, 20.159) | **$2.14 \times 10^{-5}$** |
| **Dependent variable: lnFG** | | | | | | | | | | | | |
| **Stratum *** | **n** | **β (s.e.)** | ***p*** | **n** | **β (s.e.)** | ***p*** | **n** | **β (s.e.)** | ***p*** | **n** | **β (s.e.)** | ***p*** |
| without obesity | 546 | 0.030 (0.080) | 0.708 | 472 | 0.251 (0.082) | **0.002** | 1,162 | 0.249 (0.060) | **$3.81 \times 10^{-5}$** | ** | ** | ** |
| with obesity | 467 | 0.232 (0.105) | **0.028** | 460 | 0.127 (0.079) | 0.629 | 1,425 | 0.023 (0.062) | 0.717 | ** | ** | ** |

b. Predictor: rs37386388

| | 1990–95 Samoan cohort rs37386388 | | | 2002–03 Samoan cohort rs37386388 | | | 2010 Samoan cohort rs37386388 | | | 2006–2013 Aotearoa New Zealand cohort rs37386388 | | |
|---|---|---|---|---|---|---|---|---|---|---|---|---|
| **Dependent variable: type 2 diabetes** | | | | | | | | | | | | |
| **Stratum *** | **n** | **OR (95% CI)** | ***p*** | **n** | **OR (95% CI)** | ***p*** | **n** | **OR (95% CI)** | ***p*** | **n** | **OR (95% CI)** | ***p*** |
| without obesity | 546 | 0.403 (0.139, 1.167) | 0.093 | 527 | 0.484 (0.014, 16.738) | 0.688 | 1,265 | 0.564 (0.404, 0.789) | **$8.27 \times 10^{-4}$** | 873 | 0.619 (0.421, 0.909) | **0.014** |
| with obesity | 467 | 0.783 (0.459, 1.338) | 0.371 | 556 | 0.902 (0.608, 1.338) | 0.608 | 1,596 | 0.625 (0.500, 0.781) | **$3.82 \times 10^{-5}$** | 1,251 | 0.800 (0.623, 1.028) | 0.082 |
| **Dependent variable: lnFG** | | | | | | | | | | | | |

| Stratum * | 1990–95 Samoan cohort rs37386388 | | | 2002–03 Samoan cohort rs37386388 | | | 2010 Samoan cohort rs37386388 | | | 2006–2013 Aotearoa New Zealand cohort rs37386388 | | |
|---|---|---|---|---|---|---|---|---|---|---|---|---|
| | **n** | **β (s.e.)** | ***p*** | **n** | **β (s.e.)** | ***p*** | **n** | **β (s.e.)** | ***p*** | **n** | **β (s.e.)** | ***p*** |
| without obesity | 546 | −0.034 (0.014) | **0.019** | 472 | −0.045 (0.017) | **0.010** | 1,162 | −0.022 (0.011) | **0.044** | ** | ** | ** |
| with obesity | 467 | −0.027(0.019) | 0.159 | 460 | −0.010 (0.020) | 0.629 | 1,425 | −0.042 (0.011) | **$2.34 \times 10^{-4}$** | ** | ** | ** |

*without obesity: BMI ≤ 32 $kg/m^2$; with obesity: BMI > 32 $kg/m^2$

**Fasting glucose was not measured in the Aotearoa New Zealand cohort

**Supplementary Table S3:** Effect of lnBMI on type 2 diabetes and fasting glucose in each cohort, stratified by genotype

| | 1990–95 Samoan cohort lnBMI | | | 2002–03 Samoan cohort lnBMI | | | 2010 Samoan cohort lnBMI | | | 2006–2013 Aotearoa New Zealand cohort lnBMI | | |
|---|---|---|---|---|---|---|---|---|---|---|---|---|
| **Dependent variable: type 2 diabetes** | | | | | | | | | | | | |
| **Stratum** | **n** | **OR (95% CI)** | ***p*** | **n** | **OR (95% CI)** | ***p*** | **n** | **OR (95% CI)** | ***p*** | **n** | **OR (95% CI)** | ***p*** |
| GG | 574 | 4.371 (0.497, 38.422) | 0.184 | 643 | 3.728 (1.061, 13.097) | **0.040** | 1,513 | 4.811 (2.179, 10.621) | **$1.01 \times 10^{-4}$** | 1,381 | 9.189 (4.794, 17.614) | **$2.43 \times 10^{-11}$** |
| GA | 370 | 19.011 (1.433, 252.204) | **0.026** | 392 | 29.548 (0.006, $1.47 \times 10^{5}$) | 0.436 | 1,116 | 8.576 (0.073, $1.0 \times 10^{3}$) | 0.375 | 669 | 5.912 (2.184, 16.001) | **$4.73 \times 10^{-4}$** |
| AA | 69 | ** | ** | 48 | * | * | 232 | 1.774 (0.186, 16.928) | 0.619 | 74 | *** | *** |
| **Dependent variable: lnFG** | | | | | | | | | | | | |
| **Stratum** | **n** | **β (s.e.)** | ***p*** | **n** | **β (s.e.)** | ***p*** | **n** | **β (s.e.)** | ***p*** | **n** | **β (s.e.)** | ***p*** |
| GG | 574 | 0.140 (0.061) | **0.021** | 537 | 0.223 (0.049) | **$4.58 \times 10^{-6}$** | 1,331 | 0.296 (0.037) | **$5.73 \times 10^{-16}$** | **** | **** | **** |
| GA | 370 | 0.148 (0.071) | **0.038** | 353 | 0.263 (0.068) | **$1.17 \times 10^{-4}$** | 1,036 | 0.219 (0.046) | **$2.08 \times 10^{-6}$** | **** | **** | **** |
| AA | 69 | 0.064 (0.121) | 0.597 | 42 | 0.383 (0.126) | **0.002** | 220 | 0.103 (0.088) | 0.243 | **** | **** | **** |

*Too few type 2 diabetes cases (n = 6) to run analysis.

**Too few type 2 diabetes cases (n = 2) to run analysis.

***Too few type 2 diabetes cases (n = 16) to run analysis.

****Fasting glucose was not measured in the Aotearoa New Zealand cohort

**Supplementary Figure S1:** Comparison of the cohort odds ratios and effect sizes in the obesity and genotype-stratified groups.

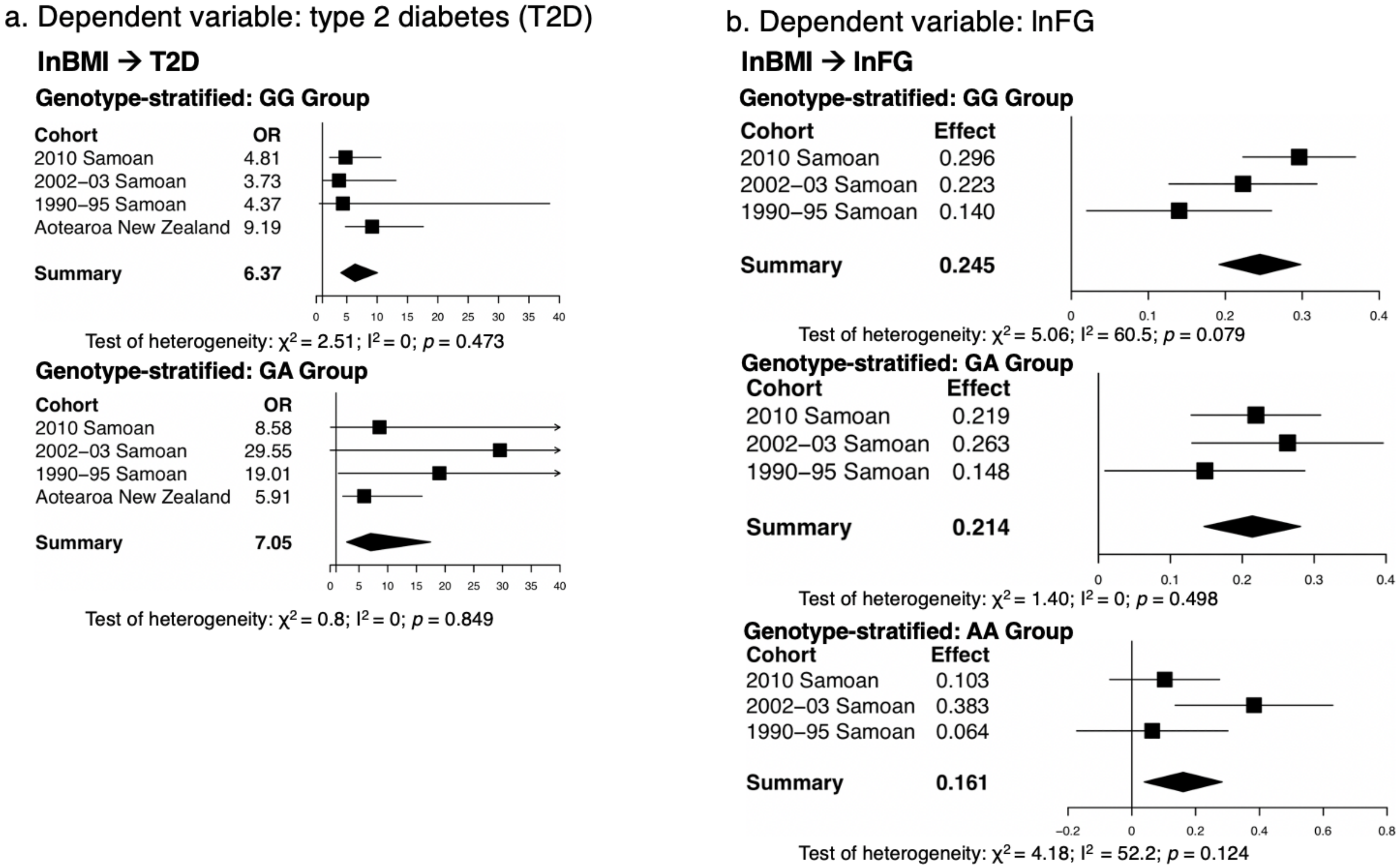

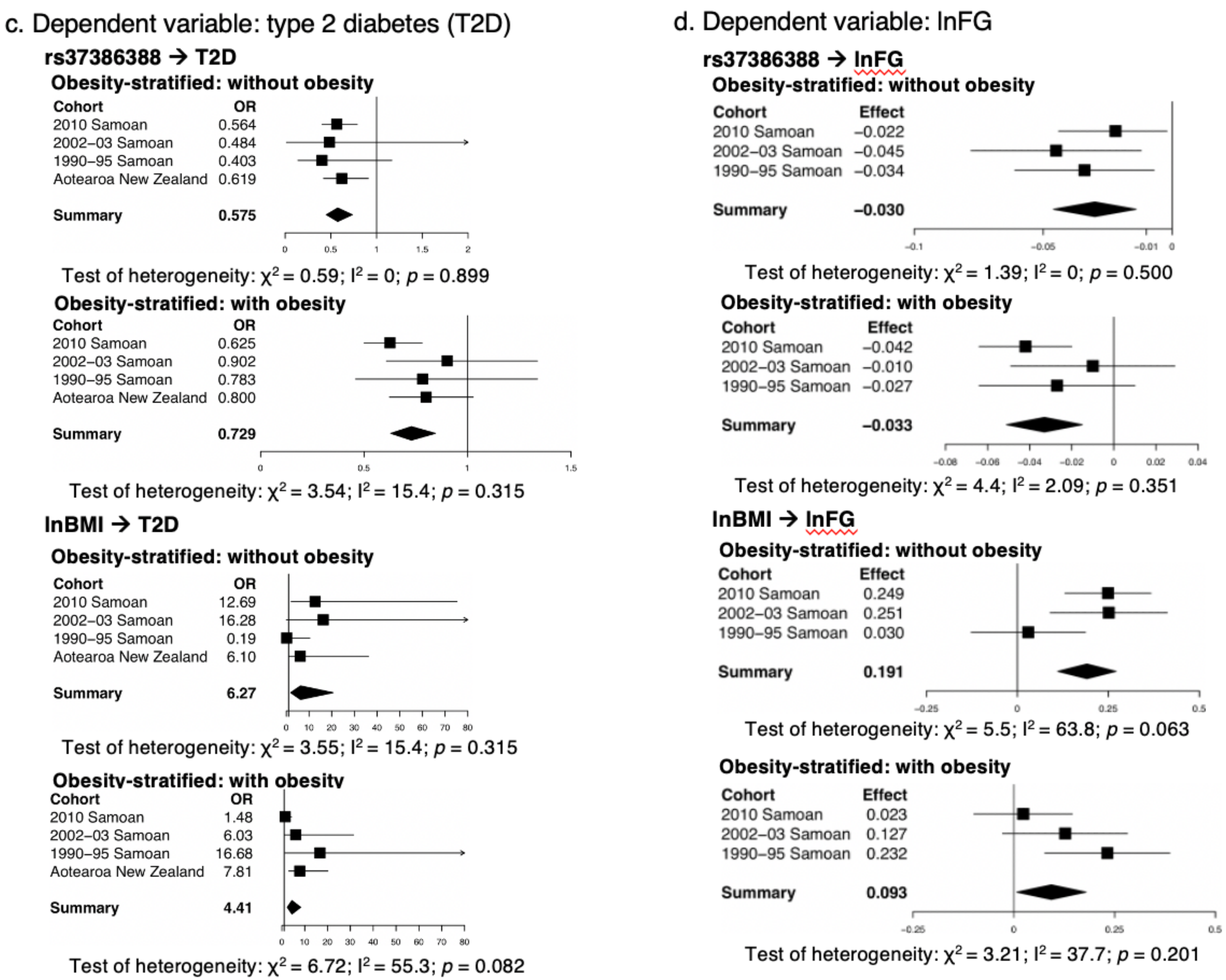
c. Dependent variable: type 2 diabetes (T2D)
rs37386388 → T2D
Obesity-stratified: without obesity
Cohort OR
2010 Samoan 0.564
2002–03 Samoan 0.484
1990–95 Samoan 0.403
Aotearoa New Zealand 0.619
Summary 0.575
0 0.5 1 1.5 2
Test of heterogeneity: χ2 = 0.59; I2 = 0; p = 0.899
Obesity-stratified: with obesity
Cohort OR
2010 Samoan 0.625
2002–03 Samoan 0.902
1990–95 Samoan 0.783
Aotearoa New Zealand 0.800
Summary 0.729
0 0.5 1 1.5
Test of heterogeneity: χ2 = 3.54; I2 = 15.4; p = 0.315
lnBMI → T2D
Obesity-stratified: without obesity
Cohort OR
2010 Samoan 12.69
2002–03 Samoan 16.28
1990–95 Samoan 0.19
Aotearoa New Zealand 6.10
Summary 6.27
0 10 20 30 40 50 60 70 80
Test of heterogeneity: χ2 = 3.55; I2 = 15.4; p = 0.315
Obesity-stratified: with obesity
Cohort OR
2010 Samoan 1.48
2002–03 Samoan 6.03
1990–95 Samoan 16.68
Aotearoa New Zealand 7.81
Summary 4.41
0 10 20 30 40 50 60 70 80
Test of heterogeneity: χ2 = 6.72; I2 = 55.3; p = 0.082
d. Dependent variable: lnFG
rs37386388 → lnFG
Obesity-stratified: without obesity
Cohort Effect
2010 Samoan –0.022
2002–03 Samoan –0.045
1990–95 Samoan –0.034
Summary –0.030
-0.1 -0.05 -0.01 0
Test of heterogeneity: χ2 = 1.39; I2 = 0; p = 0.500
Obesity-stratified: with obesity
Cohort Effect
2010 Samoan –0.042
2002–03 Samoan –0.010
1990–95 Samoan –0.027
Summary –0.033
-0.08 -0.06 -0.04 -0.02 0 0.02 0.04
Test of heterogeneity: χ2 = 4.4; I2 = 2.09; p = 0.351
lnBMI → lnFG
Obesity-stratified: without obesity
Cohort Effect
2010 Samoan 0.249
2002–03 Samoan 0.251
1990–95 Samoan 0.030
Summary 0.191
-0.25 0 0.25 0.5
Test of heterogeneity: χ2 = 5.5; I2 = 63.8; p = 0.063
Obesity-stratified: with obesity
Cohort Effect
2010 Samoan 0.023
2002–03 Samoan 0.127
1990–95 Samoan 0.232
Summary 0.093
-0.25 0 0.25 0.5
Test of heterogeneity: χ2 = 3.21; I2 = 37.7; p = 0.201

**Caption:** Forest plots for a comparison of the cohort odds ratios (OR) and 95% confidence intervals in the (a) genotype-stratified and (c) obesity-stratified models with type 2 diabetes as the dependent variable and a comparison of the cohort effect sizes and 95% confidence intervals in the (b) genotype-stratified and (d) obesity-stratified models with lnFG as the dependent variable. In the genotype-stratified analyses (a), there were too few type 2 diabetes cases to run the analyses in the 2002-03 Samoan, 1990-95 Samoan, and Aotearoa New Zealand cohorts.

**Supplementary Figure S2:** Pooled analysis Path Analysis Model with Study Cohort Variables

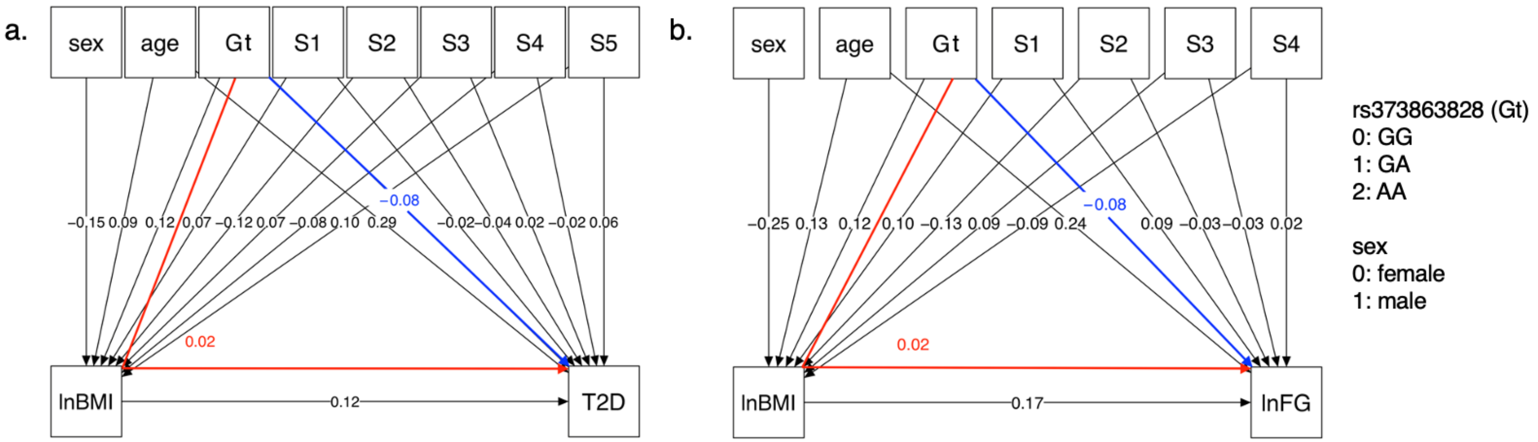

**Caption:** When study cohort variables (S1 – S5) are included in the model, the rs373863828 genotype (Gt) has both direct (blue) and indirect (red) effects on (a) type 2 diabetes (T2D) and (b) lnFG as mediated by lnBMI. For (a), the RMSEA is < 0.001 (90% CI 0.000–0.020) indicating that the path model is a very good fit. For (b), the RMSEA is < 0.064 (90% CI 0.041–0.090) indicating that the path model is a mediocre fit.
